# Correlates of prior HIV testing and schistosomiasis treatment: baseline survey findings from the “creating demand for fishermen’s schistosomiasis HIV services” (FISH) cluster-randomized trial in Mangochi, Malawi

**DOI:** 10.1101/2022.07.14.22277619

**Authors:** Geoffrey Kangogo, Donaldson F. Conserve, Sekeleghe Kayuni, Moses K. Kumwenda, Kathryn L Dovel, James Chirombo, Peter MacPherson, Elizabeth L. Corbett, Anthony Butterworth, Augustine Talumba Choko

## Abstract

**Background:** Fishing exposes fishermen to schistosomiasis-infested fresh water and concurrently through precarious livelihoods to risky sexual behaviour, rendering these two infections occupational hazards for fishermen. This study aimed to characterize the knowledge of the two conditions to obtain necessary data for a subsequent cluster randomized trial designed to investigate demand creation strategies for joint HIV-schistosomiasis service provision in fishing villages on the shores of southern Lake Malawi.

**Methods:** Enumeration of all resident fishermen in 45 clusters (fishing communities) was carried out between November 2019 and February 2020. In a baseline survey, fishermen reported their knowledge, attitudes and practices in the uptake of HIV and schistosomiasis services. Knowledge of HIV status, previous receipt of praziquantel and willingness to attend a beach clinic were modelled using random effects binomial regression, accounting for clustering.

**Results:** A total of 6,297 fishermen were surveyed from the 45 clusters with harmonic mean number of fishermen per cluster of 112 (95% CI: 97; 134). The mean age was 31.7y (SD: 11.9) and nearly 40% (2,474/6,297) could not read or write. Overall, 1,334/6,293 (21.2%) had never tested for HIV, with 64.4% (3,191/4,956) having tested in the last 12 months, and 5.9% (373/6290) taking antiretroviral therapy (ART). In adjusted analyses, being able to read and write (adjusted risk ratio [aRR: 1.91, 95% CI: 1.59-2.29, p<0.001); previous use of praziquantel (aRR: 2.00,95% CI: 1.73-2.30, p<0.001); knowing a relative or friend who died of HIV (aRR: 1.54,95% CI: 1.33-1.79, p<0.001); and being on ART (aRR: 12.93, 95% CI: 6.25-32.93, p<0.001) were associated with increased likelihood of ever testing for HIV. Only 40% (1,733/4,465) had received praziquantel in the last 12 months. Every additional year of age was associated with 1% decreased likelihood of having taken praziquantel in the last 12 months (aRR: 0.99, 95% CI: 0.98-0.99, p<0.001). However, recent HIV testing increased the likelihood of taking praziquantel by over 2-fold (aRR 2.24, 95% CI: 1.93-2.62, p<0.001). Willingness to attend a mobile beach clinic offering integrated HIV and schistosomiasis services was extremely high at 99.0% (6,224/6,284).

**Conclusion:** In a setting with an underlying high prevalence of both HIV and schistosomiasis, we found low knowledge of HIV status and low utilization of free schistosomiasis treatment. Among fishermen who accessed HIV services, there was a very high likelihood of taking praziquantel suggesting that integrated service delivery may lead to good coverage.

**Trial registration:** This trial is registered in the ISRCTN registry: ISRCTN14354324; date of registration: 05 October 2020

## Introduction

HIV and human schistosomiasis prevalence remain unacceptably high along the Great Rift Valley in Africa especially on lakeshores, due in part to complex behavioural patterns including fish-for-sex exchanges, alcoholism, condomless sex and poor access to health services(1, 2). Fish for sex trade occurs when female fish traders exchange sex with fishermen to get access to fish or to obtain competitive market prices(2). For example, fishing communities in Malawi have a higher HIV prevalence of up to 15.3% than the national prevalence of 8.8%(3). Areas of high HIV prevalence frequently overlap with highly-localised lakeshore communities where schistosomiasis prevalence is high, with urine egg-positivity in children ranging between 1%-50% in endemic areas(4). Fishing exposes fishermen to schistosomiasis-infested fresh water; concurrently, fishing exposes fishermen to risky sexual behaviour, rendering these two infections occupational hazards for fishermen.

As well as strong geographical correlation between HIV and schistosomiasis prevalence in many African countries, there are strong bidirectional assocations between the two diseases at the individual level(4). In Zambia serological evidence of previous infection with schistosomiasis was associated with an increased rates of onward transmission of HIV from co-infected men to women, and additionally increased susceptibility to HIV in HIV-negative women (4, 5). Moreover, both HIV and schistosomiasis are likely to share common risk factors and sequelae, including cervical cancer, and increased risk of infection with *Chlamydia trachomatis* and *Neisseria gonorrhoeae*(6, 7). Because many groups affected by HIV and schistosomasis have limited access to testing, health care and preventive services, an integrated approach for these marginisalised communities is required. For example, Engels et al 2020 proposed an integrated approach to HIV, schistosomiasis and cervical cancer services targeting women in Africa as an example of joint efforts(8). There is a dearth of empirical studies examining the impact of integrated schistosomiaisis and HIV service provision with only mathematical models available that suggest that such interventions could be effective and cost-effective (9, 10).

The “creating demand for fishermen’s schistosomiasis and HIV services (FISH)” cluster-randomized trial (CRT) aims to deliver HIV testing –including oral HIV self-testing (HIVST) –and *Schistosoma haematobium* services involving field microscopy and presumptive treatment i.e. treatment of all fishermen who attend the clinic in Mangochi, Malawi(11). Here we report on knowledge of HIV, previous uptake of schistosomiasis treatment and factors associated with prior HIV testing and treatment for schistosomiasis using the baseline data from the FISH CRT. We hypothesized that, as fishing communities are marginalised and have poor access to health care, knowlege of HIV and schistosomiaisis and access to severices would be low.

## Methods

### Study design

This was a cross-sectional baseline survey conducted between November 2019 and February 2020 prior to the implementation of a community cluster-randomized trial (CRT) (11). The trial’s main aim is to investigate the effect of using peer-educators, with or without HIV self-test kits, on increasing the uptake of services for schistosomiasis and HIV by adult fishermen.

### Setting

Preceding the baseline survey we undertook a formative social and physical mapping exercise on the shores of Mangochi, a district in the southeastern region of Malawi which borders Lake Malawi and Mozambique. On the southern lakeshore of Malawi, fishing is predominantly undertaken by men through small boats of up to 10 fishermen. We defined clusters to be landing sites, i.e. fishing communities on the lakeshore with at least 100 fishermen. Fishing communities were located using Google Earth maps together with local residents, and circumferential walks with global positioning satellite (GPS) devices were done to demarcate trial cluster boundaries. Community and peer leaders (fishermen representatives) were engaged to agree on the cluster boundaries, and to introduce the baseline survey and the subsequent CRT.

### Participants and procedures

The baseline survey set out to investigate knowledge, attitudes and previous testing and treatment behaviour of fishermen in southern Malawi fishing communities. Within each cluster peer leaders provided a list of fishing boats operating in the cluster as well as a list of fishermen operating in the listed boats.To be eligible for the survey, fishermen needed to: be 18 years old or over; have a fixed residence in a trial cluster; and consent to participate. We excluded fishermen who were not permanent residents of the fishing community or were underage (<18y).

Fishermen completed a face-to-face interview on tablets running open data kit (ODK) questionnaires. A total of seven interviewers were involved in the data collection. The interviewers were selected on the basis of being from within the Mangochi district to ensure quality of responses due to familiarity with the local dialect spoken in this part of the district. Interviews were conducted on the lakeshore at a private space chosen by the fishermen. All outcomes were measured via self-reports by the fishermen.

### Outcomes

Three main fisherman-level outcomes were examined in this analysis: proportion reported to have ever tested for HIV; proportion reported having tested for HIV in the last 12 months; and proportion who reported having taken praziquantel in the preceding 12 months. Explanatory variables included: age, literacy; educational attainment; male circumcision status; self-rated general health status; and knowing someone who died of HIV. Knowledge of HIV status was examined as an explanatory variable for the taking praziquantel outcome and vice versa, being indicative of health-seeking behaviour by the participant. We additionally investigated fishermens’ willingness to attend a beach clinic offering integrated HIV and schistosomiasis services, measured by asking “Would you be wiling to attend a beach clinic offering both HIV and schistosomiaisis services?”

### Statistical methods

The sample size for the survey was determined by the total number of fishermen in the clusters of the CRT. Assuming conservatively that 44% of fishermen had never tested for HIV(12) then for the sample proportion to be estimated to within +/-0.02 (2%) using the 95% confidence level, a sample of 2,367 fishermen would be required. This sample size would also be adequate to address the outcome of taking praziquantel in the last 12 months with precision of +/- 1.5%(13).

Analysis used R version 4.1.3 (R Core Data Team(14)) with p<0.05 indicating statistical significance. Frequencies were computed for categorical variables while mean and standard deviation (SD) or median and inter quartile range were computed for continuous variables that were normally distributed or skewed on visual inspection, respectively. To estimate knowledge of HIV status and exposure to praziquantel use we constructed random-effects binomial regression models, with standard errors adjusted to account for clustering. In multivariable analysis, we calculated adjusted odds ratios and 95% confidence intervals to compare between levels of explanatory variables.

## Results

A total of 6,297 participants in 45 clusters completed the baseline survey. The harmonic mean cluster size was 112 fishermen (range: 34; 245). Mean age of fishermen was 31.7 (SD: 11.9) years, with married individuals accounting for 3,924 (62.4%) (Table 1). More than half of participants were able to read and write 3819 (60.7%), but only 3616 (57.5%) had completed primary school education. Less than half of participants (40.3%) reported knowing a relative or friend who died of HIV (n=2,428) while more than half self-rated their health as good (3996/6297, 63.5%) or excellent (1321/6297, 21.0%).

**Table 1:** Demographic characteristics.

| Variable | Characteristic | Overall (N=6,297) |
| --- | --- | --- |
| Age (years) | mean (SD) | 31.7 (11.9) |
| Marital status (%) | Divorced | 279 (4.4) |
|  | Separated | 297 (4.7) |
|  | Widowed | 33 (0.5) |
|  | Never married | 1,758 (27.9) |
|  | Married | 3,924 (62.4) |
| Able to read and write? (%) | No | 2,473 (39.3) |
|  | Yes | 3,819 (60.7) |
| Highest education level attained (%) | Never been to school | 1,356 (21.5) |
|  | Primary | 3,616 (57.5) |
|  | Secondary no MSCE | 1,021 (16.2) |
|  | Secondary with MSCE | 257 (4.1) |
|  | Tertiary | 43 (0.7) |
| Tested for HIV before? (%) | No | 1,334 (21.2) |
|  | Yes | 4,959 (78.8) |
| Tested for HIV in the last 12 months? (%) | No | 1,765 (35.6) |
|  | Yes | 3,191 (64.4) |
| Currently on ART? (%) | No | 5,917 (94.1) |
|  | Yes | 373 (5.9) |
| Voluntary medical male circumcision status (%) | No | 1,389 (22.1) |
|  | Yes | 1,085 (17.2) |
|  | Local or traditional | 3,817 (60.7) |
| Ever taken praziquantel for schistosomiasis? (%) | No | 1,816 (28.9) |
|  | Yes | 4,472 (71.1) |
| Taken praziquantel for schistosomiasis in last 12 months? (%) | No | 2,732 (61.2) |
|  | Yes | 1,733 (38.8) |
| Know relative or friend who died of HIV? (%) | No | 3,599 (59.7) |
|  | Yes | 2,428 (40.3) |
| Self-rated general health (%) | Excellent | 1,321 (21.0) |
|  | Good | 3,996 (63.5) |
|  | Fair | 584 (9.3) |
|  | Poor | 393 (6.2) |
| Willing to attend a beach clinic offering HIV services? | No | 34 (0.5) |
|  | Yes | 6,255 (99.5) |
| Willing to attend a beach clinic offering schistosomiasis services? (%) | No | 13 (0.2) |
|  | Yes | 6,271 (99.8) |
| Willing to attend a beach clinic offering both HIV and schistosomiasis services? (%) | No | 60 (1.0) |
|  | Yes | 6,224 (99.0) |

SD: standard deviation; MSCE: Malawi school certificate of education; ART: antiretroviral therapy

Most partcipants (4,959, 78.8%) reported that they had previous tested for HIV; 373 (5.9%) reported taking antiretroviral therapy (ART). Overall, 71.1% (4,472) reported having taken praziquantel for schistosomiasis, with only 38.8% (n=1,733) receiving a treatment course in the preceding 12 months. Overall, most (99%, 6224/6297) of the participants reported they would be willing to attend a beach clinic offering services for HIV or schistosomiasis separately or combined.

There was considerable variation in the reported lifetime testing for HIV across the 45 clusters (Figure 1), with estimates ranging from 55.0% (95% CI: 41.6%; 67.9%) to 94.5% (95% CI: 90.9%; 96.8%). The cluster-level harmonic mean proportion was 77.2% (95% CI: 74.6; 79.9%). Similarly, we observed considerable variation in the reported recent (last 12 months) testing for HIV across the 45 clusters for fishermen who had tested for HIV before. Estimates ranged from 44.1% (95% CI: 31.2%; 51.8%) to 86.4% (95% CI: 75.0%; 93.0%). The cluster-level harmonic mean proportion was 77.2% (95% CI: 74.6; 79.9%).

**Figure 1:**
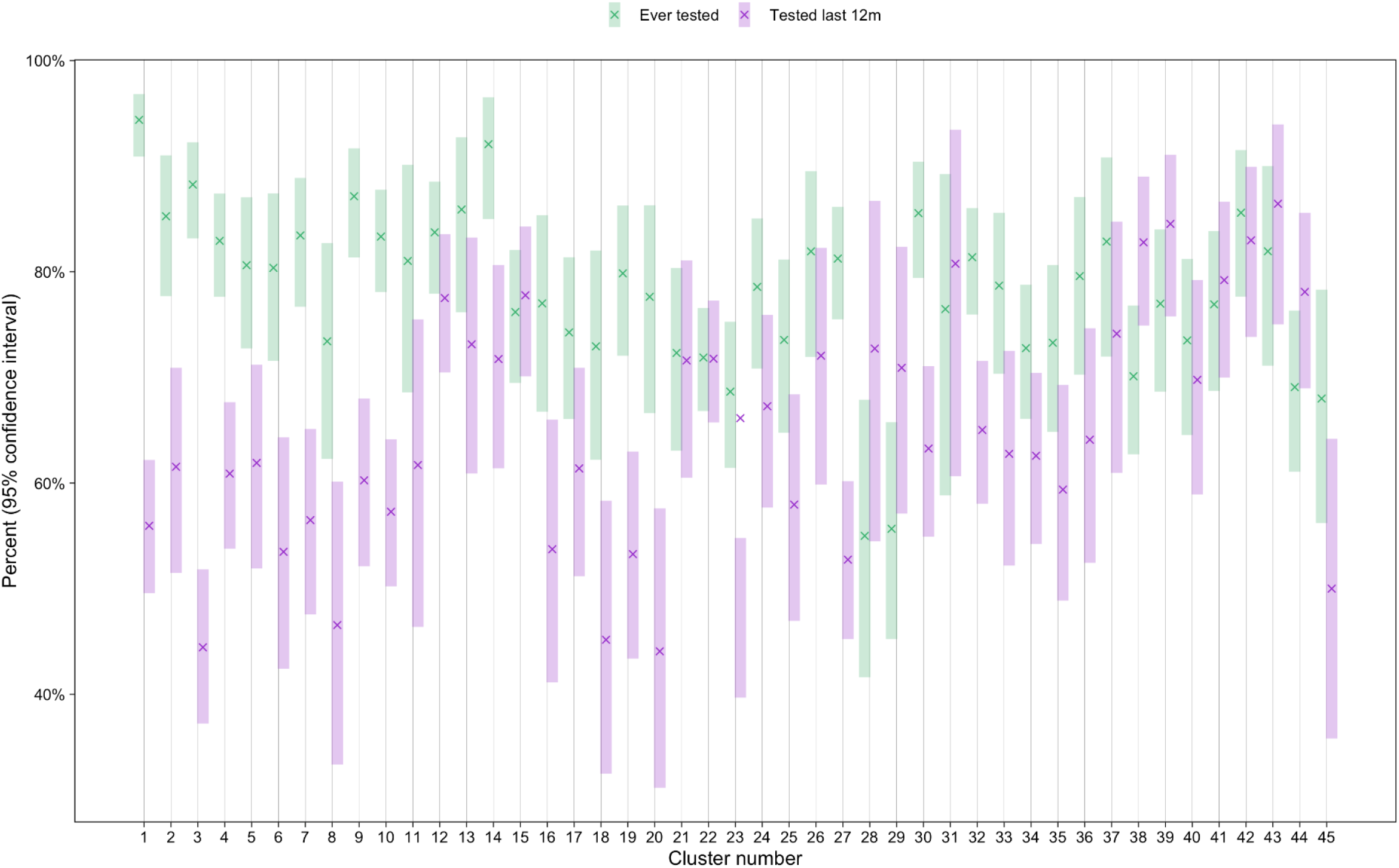
Variation in lifetime and recent HIV testing history. The variation in lifetime and recent (within last 12 months) HIV testing history revealed that most of the 45 clusters had a higher proportion threshold of over 80% of fishermen who have ever tested for HIV. In addition, two fishermen clusters (28,29) had proportions of ever tested for HIV below 70% while cluster one and 14 had their proportion peaks above 90% (Figure 1).

Table 2 shows adjusted risk ratios (aRR) for associations between explanatory variables and reported ever testing for HIV. Fishermen who could read and write, were significantly more likely to report ever testing for HIV, compared to those who could not read or write (aRR=1.91, 95% CI: 1.43; 2.30, p <0.001). Fishermen who had undergone voluntary male medical circumcision (VMMC) had 81% increased likelihood of reporting ever testing for HIV compared to those who had not undergone VMMC (aRR 1.81, 95% CI: 1.43; 2.30, p <0.001). The likelihood of reporting ever being tested for HIV increased 2-times for fishermen who had ever previously taken praziquantel compared to fishermen who had never taken praziquantel for schistosomiasis treatment (aRR 2.00, 95% CI: 1.73; 2.30, p <0.001).

**Table 2:** Association with testing for HIV in the last 12 months: unadjusted and adjusted risk ratios.

| Variable | Characteristic | Uadjusted |  |  | p-value | Adjusted |  |  | p-value |
| --- | --- | --- | --- | --- | --- | --- | --- | --- | --- |
|  |  | RR | 95% CI |  |  | aRR | 95% CI |  |  |
| Age (years) | Every additional year | 1.02 | 1.02 | 1.03 | <0.001 | 1.00 | 0.99 | 1.00 | 0.368 |
| Able to read and write? | No | 1.00 | 1.00 |  |  | 1.00 |  |  |  |
|  | Yes | 2.17 | 1.92 | 2.45 | <0.001 | 1.91 | 1.59 | 2.29 | <0.001 |
| Voluntary medical male circumcision status | No | 1.00 |  |  |  | 1.00 |  |  |  |
|  | Yes | 1.47 | 1.18 | 1.83 | 0.001 | 1.81 | 1.43 | 2.30 | <0.001 |
|  | Local or traditional | 0.72 | 0.61 | 0.84 | <0.001 | 1.02 | 0.86 | 1.21 | 0.821 |
| Ever taken praziquantel for schistosomiasis? | No | 1.00 |  |  |  | 1.00 |  |  |  |
|  | Yes | 1.98 | 1.74 | 2.24 | <0.001 | 2.00 | 1.73 | 2.30 | <0.001 |
| Know relative or friend who died of HIV? | No | 1.00 |  |  |  | 1.00 |  |  |  |
|  | Yes | 2.19 | 1.91 | 2.51 | <0.001 | 1.54 | 1.33 | 1.79 | <0.001 |
| Highest education level attained | Never been to school | 1.00 |  |  |  | 1.00 |  |  |  |
|  | Primary | 1.45 | 1.26 | 1.68 | <0.001 | 0.91 | 0.75 | 1.10 | 0.326 |
|  | Secondary no MSCE | 2.58 | 2.08 | 3.20 | <0.001 | 1.38 | 1.03 | 1.86 | 0.031 |
|  | Secondary with MSCE | 5.04 | 3.20 | 8.43 | <0.001 | 2.21 | 1.32 | 3.86 | 0.004 |
|  | Tertiary | 3.93 | 1.57 | 13.15 | 0.010 | 1.69 | 0.64 | 5.87 | 0.340 |
| Currently on ART? | No | 1.00 |  |  |  | 1.00 |  |  |  |
|  | Yes | 17.70 | 8.64 | 44.85 | <0.001 | 12.93 | 6.25 | 32.93 | <0.001 |
| Marital status | Divorced | 1.00 |  |  |  | 1.00 |  |  |  |
|  | Separated | 1.15 | 0.77 | 1.74 | 0.492 | 1.05 | 0.68 | 1.62 | 0.842 |
|  | Widowed | 1.00 | 0.43 | 2.59 | 0.993 | 1.33 | 0.52 | 3.89 | 0.569 |
|  | Never married | 0.51 | 0.38 | 0.69 | <0.001 | 0.40 | 0.28 | 0.56 | <0.001 |
|  | Married | 1.46 | 1.07 | 1.96 | 0.013 | 1.37 | 0.98 | 1.87 | 0.057 |
| Willing to attend a beach clinic offering both HIV and schistosomiasis services? | No | 1.00 |  |  |  | 1.00 |  |  |  |
|  | Yes | 1.24 | 0.67 | 2.18 | 0.472 | 1.45 | 0.73 | 2.73 | 0.266 |

aRR: adjusted risk ratio; CI: confidence interval; MSCE: Malawi school certificate of education; ART: antiretroviral therapy

The overall harmonic mean prevalence of reported praziquantel use in the last 12 months was 33.9% (95% CI: 30.4%; 38.4%) with notable variation (Figure 2). The range in the proportion of fishermen reporting previous exposure to praziquantel was 15.6% (95% CI: 8.3%; 25.6%) to 79.6% (95% CI: 70.5%; 86.9%). Every additional year of age was associated with a 1% decrease in the likelihood of having taken praziquantel in the last 12 months (aRR 0.99 (95% CI: 0.98; 0.99, p<0.001) – Table 3. Being tested for HIV in the preceding 12-months increased the likelihood of recent use (last 12-months) of praziquantel compared to fishermen who did not report having had an HIV test in the last 12 months (aRR 2.24, 95% CI: 1.93; 2.62, p<0.001).

**Table 3:**
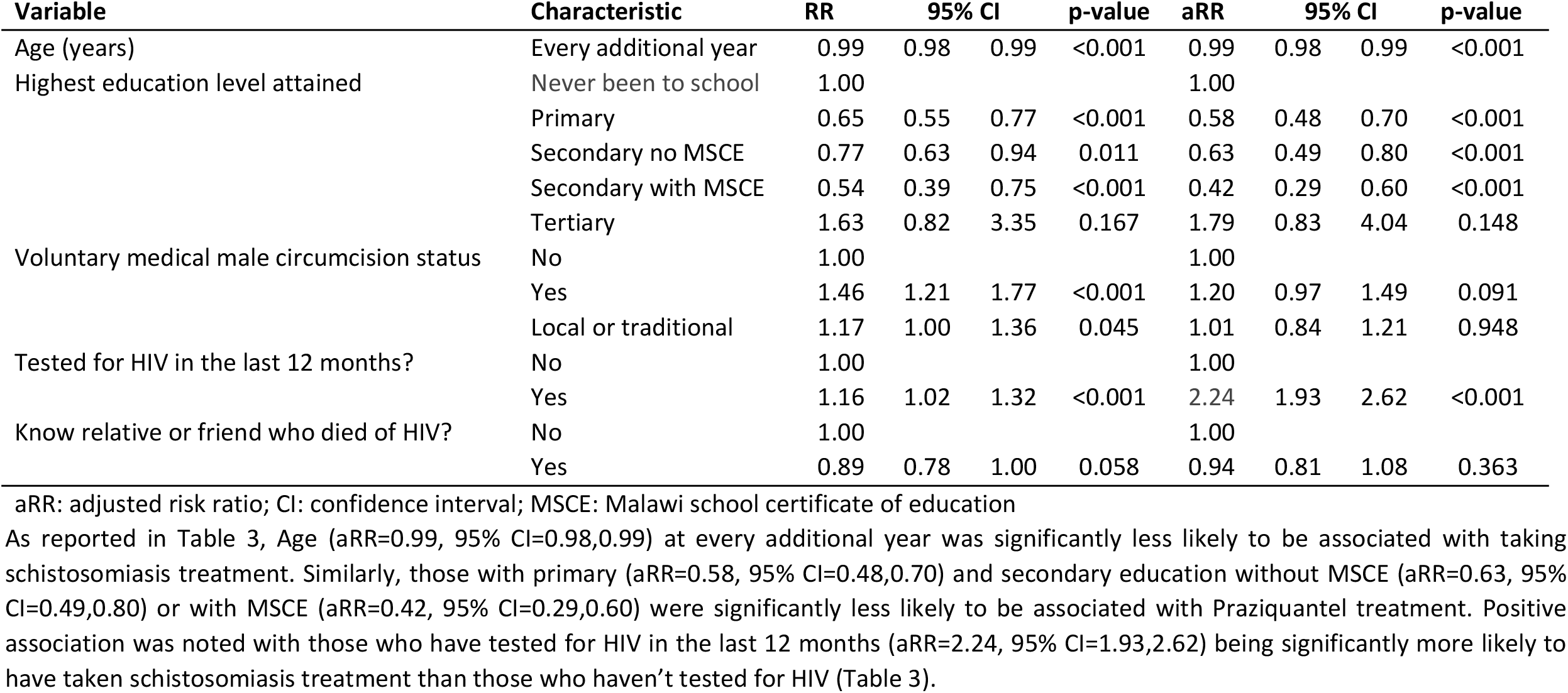
Unadjusted and adjusted associations with taking schistosomiasis treatment in the last 12 months.

**Figure 2:**
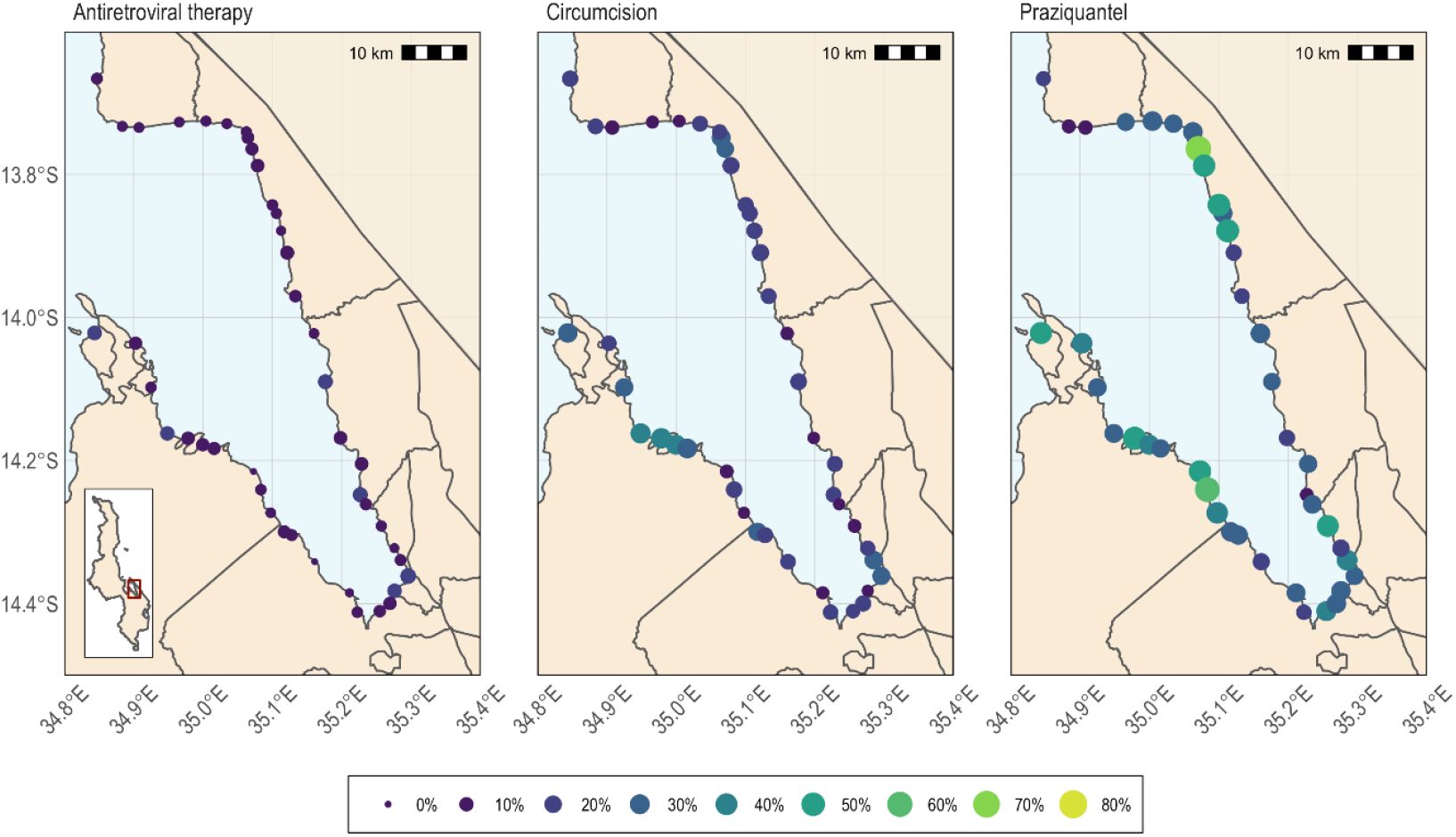
Spatial distribution of antiretroviral therap use, circumcision, and previous praziquantel use among fishermen across study clusters. The spatial distribution of Praziquantel among the fishing communities living along the shores of Lake Malawi indicating variation in prevalence from less than 10% to more than 50% (Figure 2).

Willingness to attend a beach clinic offering both HIV and schistosomiasis testing was extremely high with 99.0% of indicating they would visit such a clinic (6,224/6297) (Table 1).

## Discussion

### Key results

The main findings from this study included low utilization of free HIV services and schistosomiasis treatment (Praziquantel) among a high risk population of fishermen on the shores of Lake Malawi, (indicating that service integration is likely to increase uptake of both services), and extremely high willingness to attend beach clinics. The literature identifies mobility among fishermen, inconvenient offer of health services where there is mistiming, long distances to health facilities and stigma as key factors that contribute to low utilization(15-17). Furthermore, limited information to the fisherfolks on the significance of early testing and subsequent interventions using the approved medications is also a key contributor to low utilization of health services in this population(18). Consequently, individual, peer and societal attitudes towards drugs used to treat schistosomiasis and ART also lead to increased misinformation and heighten refusal of free drugs(19, 20).

Malawi implements twice a year mass drug administration of praziquantel among the fishing communities(21). Our data showed that surveyed fisherfolk who had received praziqualntel were more likely to have tested for HIV, and vice-versa. This is an indicator of high willingness to receive either intervention and potentially indicates acceptability of integrated HIV and schistosomiasis services. A call for such integration supports the United Nations AIDS (UNAIDS) report of 2019 which advocates for similar approaches among women (22). It is therefore plausible to develop specific interventions for HIV and schistosomiasis aimed for programmatic integration, using existing opportunities to reach fishermen and other populations at risk throughout their life course(8). The benefits of such an integrated approach should be straightforward to document the increasing uptake of HIV testing and distribution of the drug praziquantel. Elsewhere, integrated approaches have been successful in the management of female genital schistosomiasis, HIV and cervical cancer as well as successfully strengthening existing health programmes(8).

Contrary to the strongly held opinion that fishermen are unwilling to utilize health services, we found extremely high willingness to attend a mobile beach clinic when offered hypothetically among the surveyed fishermen. The key difference was that we made it clear that such a beach clinic would firstly be made available right on the lakeshore to provide convenience and secondly that fishermen would be able to access it during a wide range of hours. Such a flexible approach in service delivery has been shown in various studies and programmes to increase the uptake of services dramatically, especially for men(23).

The main strength of this study is that we showed promising results on potential significant uptake of integrated HIV and schistosomiasis services among the fishing population in Mangochi, Malawi as this will inform demand creation efforts.

The main limitation of this study was that it gives a snapshot of the baseline prevalence of the two hazards, but does not measure the impact of the services being rendered in the beach clinics. Self-reporting of HIV testing and uptake of the schistosomiasis drug praziquantel might have contributed to information bias. Given that all outcomes were self-reported there is likelihood of information bias. HIV and schistosomiasis services are free of charge and fishermen know that they are a priority population. Thus, social desirability bias would have been likely whereby fishermen may have reported that they tested for HIV or took Praziquantel when in fact they had not done so.

The study findings are generalizable to Malawi fishermen over 18 years of age and the large sample size was critical in reducing random errors. However, following this baseline study, there will be a cluster-randomized trial which will enhance both the internal and external validity of the study outcomes.

## Conclusions

In summary, we found that fishermen who accessed HIV services had a high likelihood of having previously taken the schistosomiasis drug praziquantel providing evidence that integrated service delivery may lead to increase coverage. We observed that populations in settings with a high prevalence of both HIV and schistosomiasis had low knowledge of their HIV status and low utilization of free schistosomiasis treatment. Nearly 99% of fishermen studied expressed willingness to receive both HIV and Schistosomiasis services if a mobile beach clinic is made available.

## Data Availability

https://datacompass.lshtm.ac.uk/

## Bibliography

1. MacPherson EE, Phiri M, Sadalaki J, Nyongopa V, Desmond N, Mwapasa V, et al. Sex, power, marginalisation and HIV amongst young fishermen in Malawi: Exploring intersecting inequalities. Soc Sci Med. 2020;266:113429.

2. MacPherson EE, Sadalaki J, Njoloma M, Nyongopa V, Nkhwazi L, Mwapasa V, et al. Transactional sex and HIV: understanding the gendered structural drivers of HIV in fishing communities in Southern Malawi. J Int AIDS Soc. 2012;15 Suppl 1(Suppl 1):1–9.

3. National Statistical Office/Malawi, ICF. Malawi Demographic and Health Survey 2015-16. Zomba, Malawi: National Statistical Office and ICF; 2017.

4. Kayuni S, Lampiao F, Makaula P, Juziwelo L, Lacourse EJ, Reinhard-Rupp J, et al. A systematic review with epidemiological update of male genital schistosomiasis (MGS): A call for integrated case management across the health system in sub-Saharan Africa. Parasite Epidemiol Control. 2019;4:e00077.

5. Wall KM, Kilembe W, Vwalika B, Dinh C, Livingston P, Lee YM, et al. Schistosomiasis is associated with incident HIV transmission and death in Zambia. PLoS neglected tropical diseases. 2018;12(12):e0006902.

6. Sturt AS, Webb EL, Patterson C, Phiri CR, Mweene T, Kjetland EF, et al. Cervicovaginal Immune Activation in Zambian Women With Female Genital Schistosomiasis. Frontiers in immunology. 2021;12:620657.

7. Yirenya-Tawiah D, Annang TN, Apea-Kubi KA, Lomo G, Mensah D, Akyeh L, et al. Chlamydia Trachomatis and Neisseria Gonorrhoeae prevalence among women of reproductive age living in urogenital schistosomiasis endemic area in Ghana. BMC research notes. 2014;7:349.

8. Engels D, Hotez PJ, Ducker C, Gyapong M, Bustinduy AL, Secor WE, et al. Integration of prevention and control measures for female genital schistosomiasis, HIV and cervical cancer. Bull World Health Organ. 2020;98(9):615–24.

9. Ndeffo Mbah ML, Gilbert JA, Galvani AP. Evaluating the potential impact of mass praziquantel administration for HIV prevention in Schistosoma haematobium high-risk communities. Epidemics. 2014;7:22–7.

10. Ndeffo Mbah ML, Kjetland EF, Atkins KE, Poolman EM, Orenstein EW, Meyers LA, et al. Cost-effectiveness of a community-based intervention for reducing the transmission of Schistosoma haematobium and HIV in Africa. Proceedings of the National Academy of Sciences of the United States of America. 2013;110(19):7952–7.

11. Conserve DF, Kayuni S, Kumwenda MK, Dovel KL, Choko AT. Assessing the efficacy of an integrated intervention to create demand for fishermen’s schistosomiasis and HIV services (FISH) in Mangochi, Malawi: Study protocol for a cluster randomized control trial. PloS one. 2022;17(1):e0262237.

12. Choko AT, Corbett EL, Stallard N, Maheswaran H, Lepine A, Johnson CC, et al. HIV self-testing alone or with additional interventions, including financial incentives, and linkage to care or prevention among male partners of antenatal care clinic attendees in Malawi: An adaptive multi-arm, multi-stage cluster randomised trial. PLoS medicine. 2019;16(1):e1002719.

13. Smith P, Morrow R H., Ross D A. Field Trials of Health Interventions: A Toolbox. 3rd Ed. 3rd ed 2015.

14. R Core team. A language and environment for statistical computing. Vienna, Austria: R Foundation for Statistical Computing; 2015.

15. Mafigiri R, Matovu JK, Makumbi FE, Ndyanabo A, Nabukalu D, Sakor M, et al. HIV prevalence and uptake of HIV/AIDS services among youths (15-24 Years) in fishing and neighboring communities of Kasensero, Rakai District, South Western Uganda. BMC public health. 2017;17(1):251.

16. Kiwanuka N, Ssetaala A, Nalutaaya A, Mpendo J, Wambuzi M, Nanvubya A, et al. High incidence of HIV-1 infection in a general population of fishing communities around Lake Victoria, Uganda. PloS one. 2014;9(5):e94932.

17. Seeley JA, Allison EH. HIV/AIDS in fishing communities: challenges to delivering antiretroviral therapy to vulnerable groups. AIDS care. 2005;17(6):688–97.

18. Toms K, Potter H, Balaba M, Parkes-Ratanshi R. Efficacy of HIV interventions in African fishing communities: A systematic review and qualitative synthesis. International Journal of Infectious Diseases. 2020;101:326–33.

19. Torres-Vitolas CA, Dhanani N, Fleming FM. Factors affecting the uptake of preventive chemotherapy treatment for schistosomiasis in Sub-Saharan Africa: A systematic review. PLOS Neglected Tropical Diseases. 2021;15(1):e0009017.

20. Sacolo H, Chimbari M, Kalinda C. Knowledge, attitudes and practices on Schistosomiasis in sub-Saharan Africa: a systematic review. BMC Infectious Diseases. 2018;18(1):46.

21. Makaula P, Kayuni SA, Mamba KC, Bongololo G, Funsanani M, Musaya J, et al. An assessment of implementation and effectiveness of mass drug administration for prevention and control of schistosomiasis and soil-transmitted helminths in selected southern Malawi districts. BMC health services research. 2022;22(1):517-.

22. UNAIDS. No more neglect. Female genital schistosomiasis and HIV. 2019.

23. Choko AT, Kumwenda MK, Johnson CC, Sakala DW, Chikalipo MC, Fielding K, et al. Acceptability of woman-delivered HIV self-testing to the male partner, and additional interventions: a qualitative study of antenatal care participants in Malawi. Journal of the International AIDS Society. 2017;20(1):21610.

